# Uncovering the Shared Genetic Components of Thyroid Disorders and Reproductive Health

**DOI:** 10.1101/2023.11.22.23298878

**Authors:** Jéssica Figuerêdo, Kristi Krebs, Natàlia Pujol-Gualdo, Toomas Haller, Urmo Võsa, Vallo Volke, Estonian Biobank research team, Health Informatics research team, Triin Laisk, Reedik Mägi

**Affiliations:** Estonian Genome Centre, Institute of Genomics, University of Tartu, Tartu, Estonia; Institute of Biomedicine and Translational Medicine, Department of Pathophysiology, University of Tartu, Tartu, Estonia; Institute of Computer Science, University of Tartu, Tartu, Estonia

## Abstract

Thyroid disorders are complex and polygenic, and thus result from the interaction of many genetic variants. Due to a central role of the thyroid hormones in the body, their dysfunction can have a significant impact on the reproductive health in men and women. Mapping the shared genetic component and relationships between thyroid and reproductive health traits will improve the understanding about the interplay between those domains. Here, a large-scale genetic analysis of thyroid traits (hyper- and hypothyroidism, and thyroid stimulating hormone levels) was conducted in up to 743,088 individuals of European ancestry from various cohorts. We evaluated genetic and phenotypic associations using genome-wide association study (GWAS) meta-analysis, gene prioritisation, genetic correlation analysis, and phenotype vs phenome-wide-association analysis. The results showed that 32% of thyroid-associated genes also had an impact on reproductive phenotypes, with the most affected functions being related to genitourinary tract issues. The study highlights the shared genetic determinants between thyroid function and reproductive health, providing evidence for the genetic pleiotropy between these traits in both men and women.

## INTRODUCTION

Complex diseases are a major health concern in humans and are characterized by their multifactorial nature, involving a combination of environmental, lifestyle, and genetic factors (Hunter, 2005). This makes it difficult to untangle contributing genetic factors. However, genomic data can be used to identify the genetic variants associated with a specific phenotype or disease (Marchini et al., 2005). While genetic association studies have advanced our understanding of the genetic basis of some complex diseases, maps of the shared genetic component and relationships between different traits are only beginning to emerge, for instance the association between thyroid dysfunction and reproductive health traits (Gaberšček et al., 2015; Kjaergaard et al., 2021).

The thyroid is an essential endocrine gland that plays a critical role in regulating human metabolism, growth, and development. The hypothalamus-pituitary-thyroid (HPT) axis synthesises and releases the hormones triiodothyronine (T3) and thyroxine (T4), which regulate a range of biological functions (Feldt-Rasmussen et al., 2021). Thyroid-stimulating hormone (TSH) and TSH receptor are the main controllers of thyroid function (Szkudlinski et al., 2002). If hormone production is unbalanced, it can result in hyperthyroidism or hypothyroidism, which are characterised by high or low thyroid hormone production, respectively. In addition to iodine intake, autoimmune diseases (e.g. Hashimoto’s disease), and environmental factors such as radiation exposure, selenium and vitamin D deficiency can also trigger these disorders (Ferrari et al., 2017; Vanderpump, 2011).

Given the central role of thyroid hormones in the body, thyroid problems can have a more systemic effect (Lillevang-Johansen et al., 2019, 2018; Sadek et al., 2017). In some cases, this is due to physiological interactions between the HPT axis and other hormonal systems, such as the hypothalamic-pituitary-gonadal (HPG) axis (Castañeda Cortés et al., 2014; Doufas and Mastorakos, 2000). Because of that interaction, thyroid dysfunction has a significant impact on reproductive hormone and transporter levels, such as sex-hormone binding globulin (SHBG) and testosterone (T) (Bjoro et al., 2000; Saran et al., 2016), and can also affect fertility, and menstrual and sexual function (Krassas and Markou, 2019; Nisar et al., 2012; Poppe et al., 2007; Quintino-Moro et al., 2014; Sengupta and Dutta, 2018).

Looking into the genetics, thyroid disorders are considered polygenic and result from the interaction of many genetic variants. A wide range of genes are known to be involved in thyroid dysfunctions, some with hyperthyroidism (*HLA-DR3*, *CTLA4*, *PTPN22*, *CD40*, *IL2RA*, and *FCRL3*) and others with hypothyroidism (*SLC5A5*, *SLC26A4*, *TG*, *TPO*, *DUOX2*, and *DUOXA2*) (Bufalo et al., 2021; Du et al., 2021; Geysels et al., 2022; Khong et al., 2016; Li et al., 2020; Peters et al., 2019; Pfarr et al., 2006; Ris-Stalpers and Bikker, 2010). However, none of the genes are specific to thyroid disorders (Cortés and Mendieta Zerón, 2019). In this sense, the main question is whether the associations between thyroid and reproductive health are purely due to the interactions between the regulatory axes, or if shared genetic causes might also play a role.

Genome-wide association studies (GWAS), in combination with post-GWAS analyses, are a powerful approach to explore shared genetic components between traits, identifying genetic variants/genes, and facilitating the mapping of pleiotropic associations and cross-trait genetic correlations (Mortezaei and Tavallaei, 2021; Uffelmann et al., 2021). Following a similar idea, phenome-wide-association study (PheWAS) gives information on which conditions are more likely to be present among individuals with the same genetic variant. Furthermore, one can expand this approach to study specific phenotypes/diagnoses (Phe-PheWAS) and their associations with many other phenotypes (Claussnitzer et al., 2020).

Here we use large-scale genetic analyses of three thyroid traits (hyperthyroidism, hypothyroidism, and measured TSH levels) in up to 743,008 European ancestry individuals to explore their associations with several reproductive health traits, both on the phenotypic and genetic level (Figure 1). We identify genes that have a pleiotropic role in thyroid and reproductive function, characterise genetic correlations between thyroid and reproductive traits, and finally use individual-level Estonian Biobank data to identify diagnoses associated with thyroid disorders in men and women.

**Figure 1.**
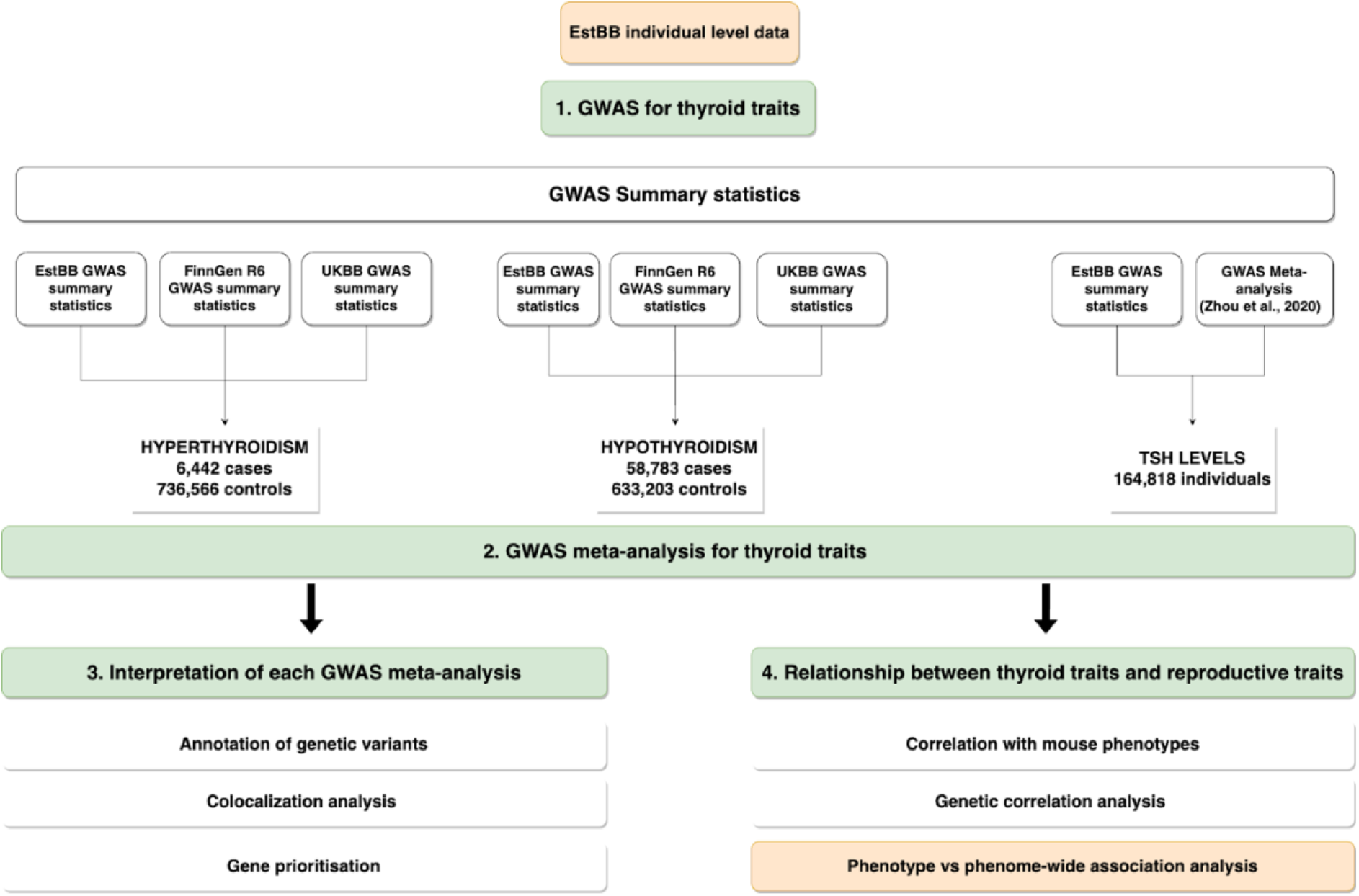
Flowchart of the study approach.

## METHODS

### Estonian Biobank cohort

Estonian Biobank (EstBB) is a population-based cohort of approximately 200,000 participants (∼20% of the Estonian adult population) with a rich variety of phenotypic and health-related information collected for each individual (Leitsalu et al., 2015). At recruitment, participants have signed a consent to allow follow-up linkage of their electronic health records (EHR), including lab measurements, thereby providing a longitudinal collection of phenotypic information. EstBB links regularly with the National Health Insurance Fund (from 2004) and other relevant registries. For every participant there is information on diagnoses in International Classification of Disease 10 (ICD-10) codes (version 2019). Briefly, we used ICD-10 codes to identify individuals with hyper- and hypothyroidism. In addition, we used data on TSH measurements that were extracted from free-text medical notes. More information about the definition of phenotypes is provided in the GWAS-EstBB data below.

### EstBB Genotype data

Genotyping of DNA samples from the EstBB was done at the Core Genotyping Lab of the Institute of Genomics, University of Tartu using the Illumina Global Screening Arrays (GSAv1.0, GSAv2.0, and GSAv2.0_EST). Altogether 200,000 samples were genotyped and then PLINK format files were created using Illumina GenomeStudio v2.0.4. During the quality control all individuals with call-rate < 95% or mismatching sex that was defined based on the heterozygosity of X chromosome and sex in the phenotype data, were excluded from the analysis. Variants were filtered by call-rate < 95% and HWE p-value < 1×10^−4^ (autosomal variants only). Variant positions were in Genome Reference Consortium Human Build 37 and all variants were changed to be from TOP strand using reference information provided by Dr. Will Rayner from the University of Oxford (https://www.well.ox.ac.uk/∼wrayner/strand/). Before imputation variants with MAF<1% and indels were removed. Prephasing was done using the Eagle v2.3 software (Loh et al., 2016) (number of conditioning haplotypes Eagle2 uses when phasing each sample was set to: --Kpbwt=20000) and imputation was carried out using Beagle v.28Sep18.793 (Browning et al., 2018; Browning and Browning, 2007) with an effective population size ne=20,000. As a reference, Estonian population specific imputation reference of 2,297 WGS samples was used (Mitt et al., 2017).

### GWAS-EstBB data

For the current study, we selected cases and controls using ICD-10 codes and drug prescription information for hyperthyroidism (4,825 cases and 143,282 controls), and for hypothyroidism (11,075 cases and 178,004 controls). Detailed information on the phenotype definition can be found in the Supplementary table S1. Controls were defined as individuals not identified as cases for the phenotypes.

For the GWAS of thyroid stimulating hormone, the measurements of Logical Observation Identifiers Names and Codes (LOINC) code 3016-3 (“Thyrotropin in Serum or Plasma”) were obtained from EHR. During the quality control we removed outliers that were outside of three standard deviations from the mean. The raw data extraction workflow can be found in the Supplementary information (Text S3) of Kurvits et al. (2023). For individuals with multiple measurements the most recent available measurement was used for GWAS analysis. TSH levels were obtained from 45,103 individuals and the levels ranged from 0 to 35.99 mIU/L, with a median of 1.62 and mean of 2.00 (sd=1.81).

We conducted sex combined GWASs for hyperthyroidism, hypothyroidism and TSH using the REGENIE software (version 3.2) (Mbatchou et al., 2021) adjusting for the first ten genotype principal components (PCs), year of birth, and sex.

### FinnGen cohort

FinnGen research project is a public-private partnership which contains 392,000 Finnish individual’s genome information combined with digital health record data. The data release R6 (https://r6.finngen.fi/) was used for the analyses presented here. FinnGen individuals were genotyped with Illumina and Affymetrix arrays (Illumina Inc., San Diego, and Thermo Fisher Scientific, Santa Clara, CA, USA). Genotype data imputation was performed using the population-specific SISu v3 imputation reference panel of 3,775 whole genomes. Disease endpoints for the biobank have been electronically defined based on nationwide registries. GWAS analyses were done using SAIGE (0.39.1), using sex, age, 10 genotype PCs and genotyping batch as covariates in the model (Kurki et al., 2022).

We downloaded GWAS summary statistics available for the endpoint “Autoimmune hyperthyroidism”, total of 204,701 individuals (1,161 cases and 203,540 controls); and for “Hypothyroidism (congenital or acquired)” which presented a total of 98,381 individuals (32,925 cases and 65,456 controls). Those endpoints were chosen because they are the broader endpoint for the specific diseases and, consequently, have the largest sample size. More information about the phenotype definitions can be found in the Supplementary table S1.

### UK Biobank cohort

UK Biobank (UKBB) is a large population-based study that has genomic and phenotype data for more than 500,000 individuals. For the meta-analysis, GWAS summary statistics were obtained via the PheWeb portal (https://pheweb.org/UKB-TOPMed/phenotypes) which contains EHR-derived ICD codes from the White British participants of the UKBB. Genotype data had been imputed using Trans-Omics for Precision Medicine (TOPMed) and GWAS analyses were conducted using SAIGE adjusting for genetic relatedness, sex, year of birth and the first four principal components. The summary statistics for "Graves’ disease" (autoimmune hyperthyroidism; PheCode 242.1) and for "Hypothyroidism" (PheCode 244) were used, with 390,200 individuals (456 cases and 389,744 controls) and 404,529 individuals (14,783 cases and 389,742 controls), respectively. More info about the phenotype definitions can be found in the Supplementary table S1.

### HUNT, MGI and ThyroidOmics Consortium

For analysis of TSH phenotype, we meta-analyzed GWAS summary statistics from EstBB, and from Zhou et al. (2020). The latter included European ancestry individuals from three projects: HUNT, MGI and ThyroidOmics Consortium, with sample sizes of 55,342 individuals, 10,085 individuals and 54,288 individuals, respectively. The summary statistics of this meta-analysis were downloaded from: http://csg.sph.umich.edu/willer/public/TSH2020/.

Nord-Trøndelag Health Study (HUNT) is a health study that has DNA samples from about 70,000 participants genotyped using Illumina HumanCoreExome v1.0 and 1.1, and imputed using Minimac3 with a merged reference panel of Haplotype Reference Consortium (HRC) (Krokstad et al., 2013).

Michigan Genomics Initiative (MGI) is a collaborative research project composed particularly by pre-surgical patients from Michigan Medicine. It has more than 85,000 participants genotyped using Illumina Infinium CoreExome-24 bead array platform and imputed by software Minimac4 (v1.0.0) with HRC and TOPMed (Zawistowski et al., 2021).

ThyroidOmics consortium is a meta-analysis dataset composed of several cohorts (up to 119,715 individuals) whose aim is to have a deep understanding of determinants and effects of thyroid diseases based on the genotype and phenotype data.

### GWAS meta-analysis

We performed a meta-analysis of GWAS summary statistics for each phenotype [hyperthyroidism (6,442 cases and 736,566 controls), hypothyroidism (58,783 cases and 633,203 controls) and TSH levels (164,818 individuals)] using the inverse of variance based fixed effect method implemented into GWAMA (Mägi and Morris, 2010).

For hyperthyroidism and hypothyroidism, we meta-analyzed GWAS summary statistics from EstBB, FinnGen and UKBB; and for TSH levels, we conducted a meta-analysis using EstBB GWAS summary statistics and the summary statistics created by Zhou et al. (2020). Genome-wide statistical significance in each analysis was defined as p<5×10^−8^.

### Annotation of GWAS signals

Annotation of GWAS signals was done using the Functional Mapping and Annotation of Genome-Wide Association Studies (FUMA) platform (v1.3.6a) (Watanabe et al., 2017). FUMA defines genomic risk loci from independent significant SNPs (Single Nucleotide Polymorphisms) by merging LD (Linkage Disequilibrium) blocks, gene mapping and in addition annotates (using ANNOVAR) all input variants also including information for variants in LD. Novel loci were defined when there was no previous association between the locus (lead variant + variants in LD) and thyroid phenotypes, using GWAS catalog and previous publications as search base. We used data on nearest genes to index variants in each genomic risk locus and SNP annotations for further down-stream analyses.

### Colocalization

For colocalization analysis between GWAS meta-analysis summary statistics and eQTL Catalog dataset (Kerimov et al., 2021), we used HyprColoc (Foley et al., 2021). We selected the Genotype Tissue Expression (GTEx v8) thyroid tissue dataset for the analysis (dataset ID: QTD000341) (THE GTEX CONSORTIUM, 2020). The p-value threshold to identify lead SNPs was set to 5×10^−8^ and the window around each SNP to test the colocalization to 1MB.

### Gene prioritisation

To determine the possible candidate causal genes at each genomic risk locus, we carried out gene prioritisation using the following criteria: 1) proximity to lead variant; 2) significant (posterior probability ≥ 0.8) colocalization between GWAS signal and gene eQTL in thyroid tissue; 3) genes containing missense variants or in high LD (r2 > 0.6) with these; and 4) genes which showed thyroid-related phenotypes in mutant mice or monogenic thyroid diseases in humans.

To select the nearest gene for each locus we used information provided by FUMA and did an additional search using the Open Target Genetics tool (Ghoussaini et al., 2021; Mountjoy et al., 2021). We prioritised protein coding genes and, if there was a conflict in gene mapping between the two sources, we preferred genes that were previously reported as being associated with thyroid traits in GWAS Catalog or had more associations in the gene prioritisation analysis. Mouse Genome Database (Blake et al., 2020) was utilised to evaluate the association between our variants and mouse phenotypes and monogenic human diseases. The model dataset present in the browser is integrated with human gene-to-disease relationships from the National Center for Biotechnology Information (NCBI) and Online Mendelian Inheritance in Man (OMIM), and human disease-to-phenotype relationships from the Human Phenotype Ontology (HPO).

### Thyroid and Reproductive health traits

To evaluate the correlation between thyroid diseases and reproductive health phenotypes, we carried out the following analyses:

#### Correlation with mouse phenotypes

Using the Mouse Genome Database, we evaluated available mouse model phenotypes to identify the association between our candidate genes and mouse phenotypes or human monogenic diseases and respective phenotypes related to reproductive health.

#### Genetic correlation analysis

This analysis was carried out for the three thyroid phenotypes using the LDScore tool implemented in Complex Trait Genetics Virtual Lab (0.4-beta) (B. Bulik-Sullivan et al., 2015; B. K. Bulik-Sullivan et al., 2015; Cuellar-Partida et al., 2019). Analyses were performed between the three generated meta-analysis datasets and: 1) all traits available in CTG browser (N=1,376); 2) GWAS summary statistics for reproductive hormones/traits (N=25) for European ancestry individuals available either in GWAS Catalog, created by Pott et al. (2019) or from personal data (Supplementary table S2). Benjamini-Hochberg FDR correction was used to identify statistically significant associations [0.05 x (p-value rank/total number of tests)].

#### Phenotype vs phenome-wide association analysis (Phe-PheWAS)

To evaluate the diseases associated with the three studied phenotypes we performed a Phe-PheWAS analysis (Carroll et al., 2014) using R software (3.6.1). We tested the associations between sex-stratified individual level data [hyperthyroidism (N men= 610 cases and 55,944 controls; N women= 4,267 cases and 88,612 controls), hypothyroidism (N men= 900 cases and 66,845 controls; N women= 10,306 cases and 112,832 controls) and for TSH levels (11,320 men and 34,230 women)] from EstBB and 2,001 ICD-10 codes using logistic regression, excluding closely related individuals (IBD>20%) and adjusting for age and 10 genotype PCs to adjust for population stratification. Bonferroni correction was applied to select statistically significant associations (number of tested ICD codes - 2,001; Bonferroni corrected p-value threshold 0.05/2,001=2.5×10^−5^).

## RESULTS

### GWAS meta-analysis of thyroid traits

We used publicly available datasets and data from the EstBB to conduct GWAS meta-analyses for hyperthyroidism, hypothyroidism, and TSH levels with the largest available sample size for these thyroid phenotypes. In total, we identified 237 genome-wide significant associations - 15 for hyperthyroidism, 141 for hypothyroidism, and 90 for TSH (Figure 2; Supplementary table S3). Eight loci were shared between the thyroid phenotypes: rs2476601 was associated with all three thyroid phenotypes, and seven loci (rs11675342, rs310755, rs2983511, rs2921053, rs9298749, rs10748781 and rs11830537) were shared between hypothyroidism and TSH levels. Based on lookups in the GWAS Catalog and previous publications, we found 47/237 to be novel in the context of thyroid biology (three for hyperthyroidism, 34 for hypothyroidism, 10 for TSH (Table 1). Three hypothyroidism lead variants (rs546039326, rs182482858 and rs76715626) showed higher effect allele frequency in EstBB and FinnGen cohorts, when compared to other European populations such as the UKBB, fold enrichments varied from 14.7 to 171.4 (Table 1). There was no significant heterogeneity across cohorts after Bonferroni multiple testing correction (heterogeneity p-value threshold - hyperthyroidism and hypothyroidism = 6.6×10^−10^; TSH = 1.2×10^−9^) (Supplementary table S3).

**Figure 2.**
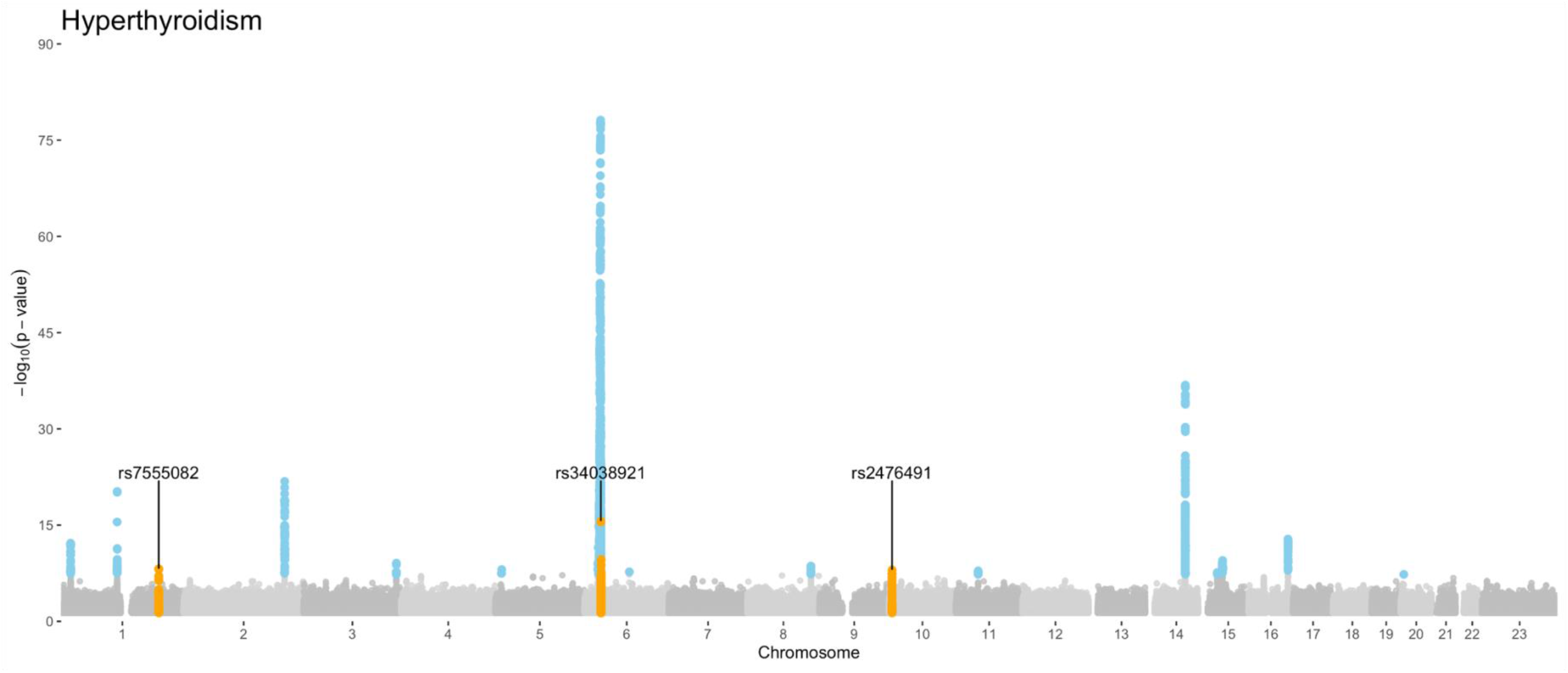
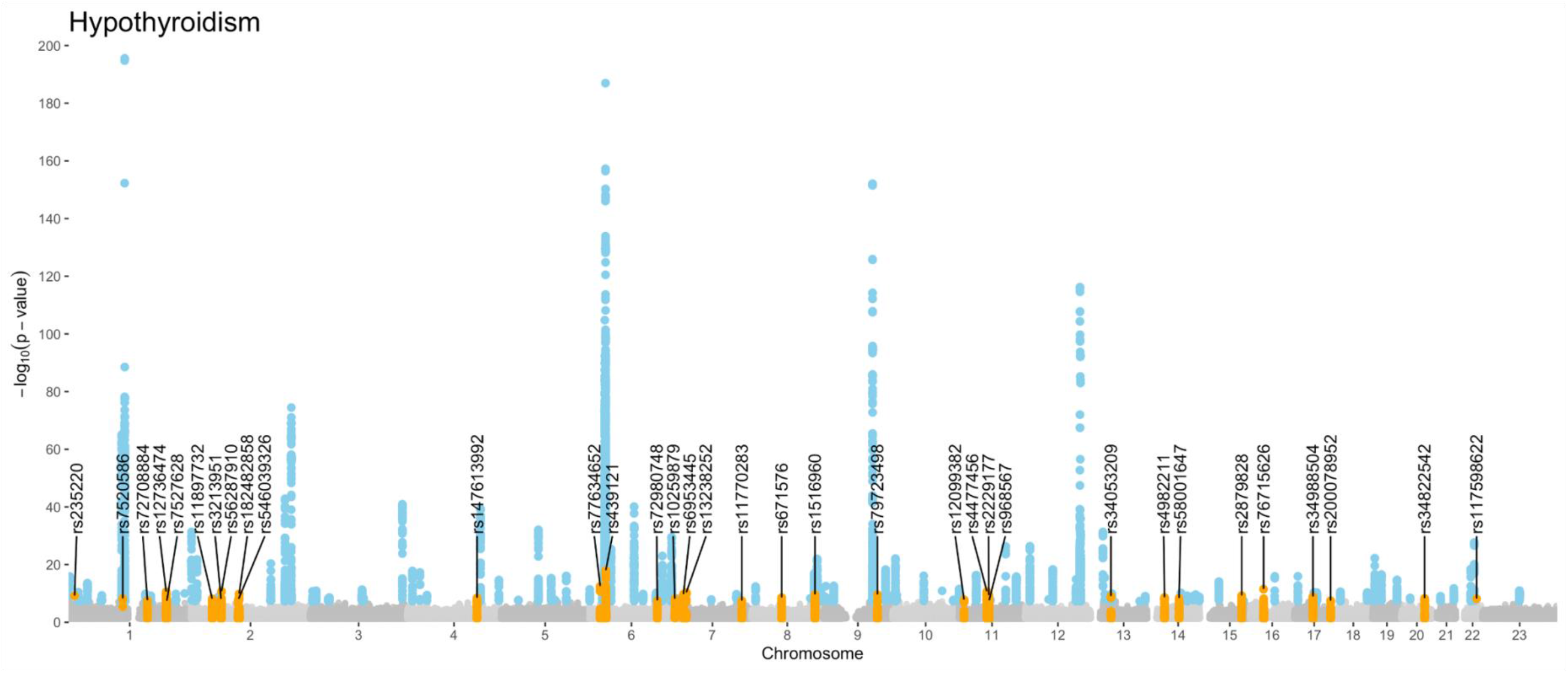
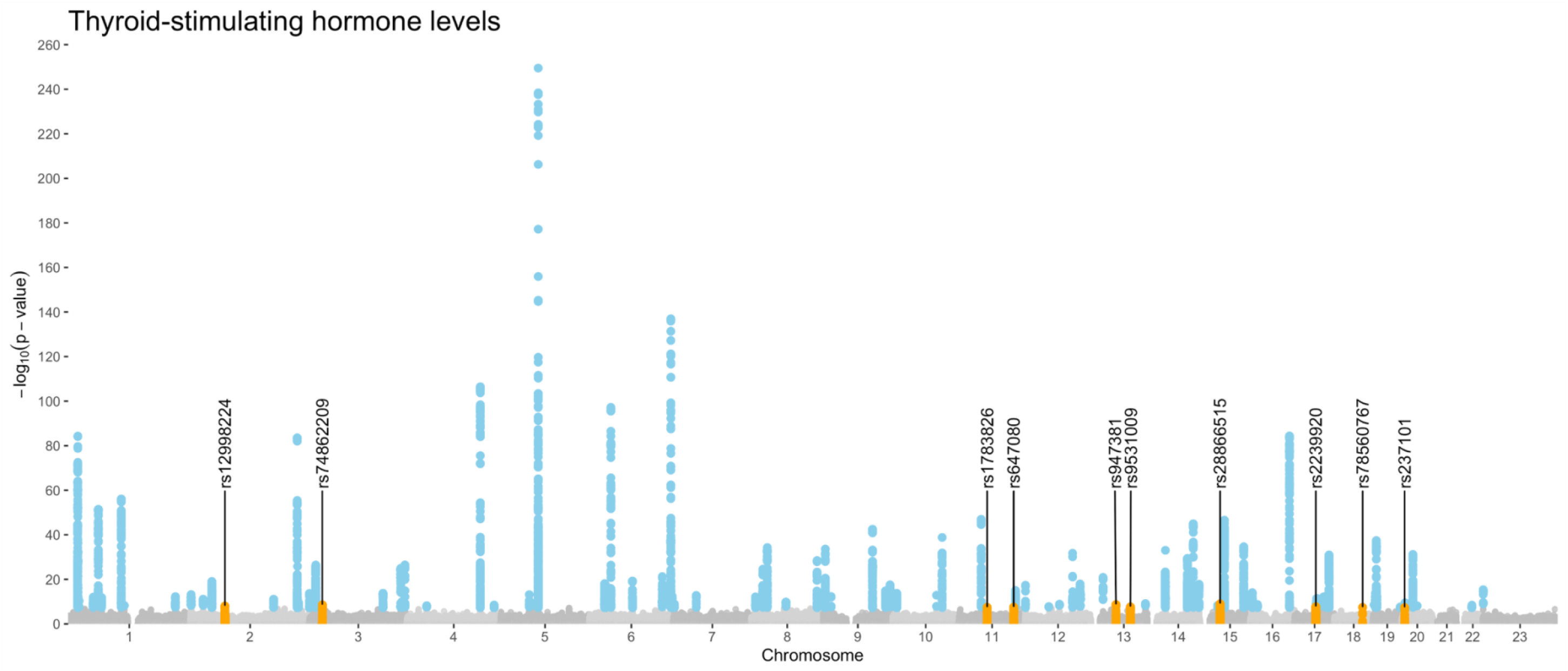
GWAS meta-analysis results for thyroid phenotypes. Manhattan plots for hyperthyroidism, hypothyroidism and TSH measurement levels meta-analysis, respectively. Genome-wide significant results are highlighted in blue (loci already known in thyroid perspective) and orange (novel loci found in this study). RS numbers for the novel loci are annotated in the figure.

**Table 1.** Summary of GWAS meta-analysis results for novel loci in hyperthyroidism, hypothyroidism and TSH levels. Chr= chromosome, EA= effect allele, NEA= non-effect allele, EAF= effect allele frequency, EstBB= Estonian Biobank, UKBB= UK Biobank, OR= odds ratio, CI= confidence interval, SE= standard error.

| NOVEL LOCI - HYPERTHYROIDISM |  |  |  |  |  |  |  |  |
| --- | --- | --- | --- | --- | --- | --- | --- | --- |
| Chr (band) | POS (hg19) | Lead variant | EA/ NEA | EAF |  |  | OR (95% CI) | P-value |
|  |  |  |  | EstBB | FinnGen | UKBB |  |  |
| chr1 (q31.3) | 198598663 | rs7555082 | G/A | 0.11 | 0.07 | 0.12 | 1.2 (1.13-1.28) | $6.9 \times 10^{-9}$ |
| chr6(p21.32) | 33195383 | rs34038921 | C/G | 0.05 | 0.05 | 0.05 | 1.41 (1.30-1.54) | $2.6 \times 10^{-16}$ |
| chr10(p15.1) | 6095410 | rs2476491 | A/T | 0.31 | 0.25 | 0.3 | 1.12 (1.08-1.17) | $1.0 \times 10^{-8}$ |
| NOVEL LOCI - HYPOTHYROIDISM |  |  |  |  |  |  |  |  |
| Chr (band) | POS (hg19) | Lead variant | EA/ NEA | EAF |  |  | OR (95% CI) | P-value |
|  |  |  |  | EstBB | FinnGen | UKBB |  |  |
| chr1(p36.22) | 12263919 | rs235220 | C/T | 0.19 | 0.21 | 0.2 | 1.06 (1.04-1.09) | $6.8 \times 10^{-10}$ |
| chr1(p13.3) | 110346147 | rs7520586 | G/T | 0.58 | 0.56 | 0.56 | 1.05 (1.03-1.07) | $6.6 \times 10^{-9}$ |
| chr1(q23.3) | 160643793 | rs72708884 | G/A | 0.07 | 0.06 | 0.05 | 1.10 (1.06-1.13) | $2.7 \times 10^{-8}$ |
| chr1(q32.1) | 198956465 | rs12736474 | A/G | 0.27 | 0.31 | 0.35 | 1.06 (1.04-1.08) | $2.8 \times 10^{-11}$ |
| chr1(q32.1) | 200259948 | rs7527628 | T/A | 0.67 | 0.69 | 0.67 | 1.05 (1.03-1.06) | $4.0 \times 10^{-8}$ |
| chr2(p21) | 43541386 | rs11897732 | A/G | 0.55 | 0.58 | 0.55 | 1.05 (1.03-1.06) | $8.2 \times 10^{-9}$ |
| chr2(p15) | 61622251 | rs3213951 | T/C | 0.32 | 0.41 | 0.33 | 1.05 (1.03-1.07) | $8.9 \times 10^{-9}$ |
| chr2(p15) | 62532084 | rs56287910 | T/C | 0.44 | 0.44 | 0.39 | 1.06 (1.04-1.07) | $1.9 \times 10^{-11}$ |
| chr2(q11.2) | 97226518 | rs546039326 | A/C | 0.01 | 0.02 | 0.00068 | 1.24 (1.15-1.34) | $9.3 \times 10^{-9}$ |
| chr2(q11.2) | 98506529 | rs182482858 | C/T | 0.01 | 0.02 | 0.00013 | 1.28 (1.19-1.39) | $1.5 \times 10^{-10}$ |
| chr4(q31.21) | 142462614 | rs147613992 | A/AC | 0.16 | 0.12 | NA | 1.09 (1.06-1.12) | $3.9 \times 10^{-9}$ |
| chr6(p22.3) | 21862343 | rs77634652 | C/G | 0.22 | 0.24 | 0.16 | 1.07 (1.05-1.10) | $3.4 \times 10^{-13}$ |
| chr6(p21.32) | 33192867 | rs439121 | A/C | 0.26 | 0.30 | 0.26 | 1.08 (1.06-1.10) | $1.1 \times 10^{-18}$ |
| chr6(q23.3) | 138150539 | rs72980748 | C/T | 0.20 | 0.20 | 0.21 | 1.06 (1.04-1.08) | $3.2 \times 10^{-8}$ |
| chr7(p22.2) | 2934386 | rs10259879 | G/A | 0.26 | 0.23 | 0.28 | 1.05 (1.03-1.07) | $8.5 \times 10^{-9}$ |
| chr7(p21.1) | 20457974 | rs6953445 | C/A | 0.65 | 0.63 | 0.65 | 1.05 (1.04-1.07) | $1.9 \times 10^{-10}$ |
| chr7(p15.2) | 26153496 | rs13238252 | G/A | 0.17 | 0.19 | 0.21 | 1.07 (1.05-1.09) | $5.2 \times 10^{-11}$ |
| chr7(q34) | 139436551 | rs11770283 | G/T | 0.22 | 0.21 | 0.19 | 1.06 (1.04-1.08) | $3.7 \times 10^{-8}$ |
| chr8(q12.1) | 61576798 | rs671576 | G/C | 0.45 | 0.48 | 0.49 | 1.05 (1.03-1.07) | $3.1 \times 10^{-9}$ |
| chr8(q24.21) | 129474882 | rs1516960 | A/T | 0.26 | 0.23 | 0.27 | 1.06 (1.04-1.08) | $2.6 \times 10^{-10}$ |
| chr9(q31.2) | 110602043 | rs79723498 | A/AC | 0.07 | 0.15 | 0.06 | 1.09 (1.06-1.12) | $2.8 \times 10^{-10}$ |
| chr11(p15.4) | 10532922 | rs12099382 | A/T | 0.05 | 0.03 | 0.01 | 1.14 (1.09-1.19) | $1.6 \times 10^{-8}$ |
| chr11(q12.1) | 57197781 | rs4477456 | G/A | 0.24 | 0.25 | 0.22 | 1.06 (1.04-1.08) | $2.2 \times 10^{-11}$ |
| chr11(q12.2) | 60893235 | rs2229177 | C/T | 0.57 | 0.6 | 0.49 | 1.05 (1.03-1.07) | $4.5 \times 10^{-10}$ |
| chr11(q12.2) | 61595564 | rs968567 | C/T | 0.13 | 0.10 | 0.19 | 1.06 (1.04-1.09) | $3.0 \times 10^{-8}$ |
| chr13(q14.11) | 41151955 | rs34053209 | C/CAG | 0.31 | 0.25 | 0.26 | 1.06 (1.04-1.07) | $9.5 \times 10^{-10}$ |
| chr14(q13.1) | 35259151 | rs4982211 | G/A | 0.13 | 0.16 | 0.1 | 1.07 (1.05-1.10) | $2.6 \times 10^{-9}$ |
| chr14(q23.3) | 64906530 | rs58001647 | T/G | 0.27 | NA | 0.26 | 1.07 (1.04-1.09) | $4.9 \times 10^{-9}$ |
| chr15(q25.3) | 85361977 | rs2879828 | G/T | 0.54 | 0.46 | 0.46 | 1.05 (1.03-1.07) | $6.1 \times 10^{-10}$ |
| chr16(p12.1) | 27398042 | rs76715626 | C/T | 0.02 | 0.06 | 0.00035 | 1.18 (1.13-1.23) | $3.4 \times 10^{-12}$ |
| chr17(q12) | 37957632 | rs34988504 | C/T | 0.06 | 0.04 | 0.03 | 1.13 (1.08-1.17) | $1.3 \times 10^{-9}$ |
| chr17(q25.1) | 73976675 | rs200078952 | ATTT/A | 0.03 | 0.05 | 0.05 | 1.11 (1.07-1.15) | $3.7 \times 10^{-8}$ |
| chr20(q13.13) | 47257755 | rs34822542 | C/T | 0.18 | 0.13 | 0.16 | 1.07 (1.04-1.09) | $5.6 \times 10^{-9}$ |
| chr22(q13.2) | 42244577 | rs117598622 | A/G | 0.01 | 0.01 | 0.01 | 1.24 (1.15-1.33) | $7.6 \times 10^{-9}$ |

| NOVEL LOCI - THYROID-STIMULATING HORMONE |  |  |  |  |  |  |  |
| --- | --- | --- | --- | --- | --- | --- | --- |
| Chr (band) | POS | Lead variant | EA/ NEA | EAF |  | BETA (SE) | P-value |
|  |  |  |  | EstBB | Meta-analysis<br>Zhou et al. (2020) |  |  |
| chr2(p13.3) | 70336418 | rs12998224 | A/C | 0.32 | 0.32 | 0.03 (0.005) | $9.1 \times 10^{-9}$ |
| chr3(p24.2) | 25596524 | rs74862209 | T/A | 0.05 | 0.06 | 0.05 (0.009) | $2.8 \times 10^{-9}$ |
| chr11(q12.1) | 57402964 | rs1783826 | G/T | 0.60 | 0.56 | 0.02 (0.004) | $3.5 \times 10^{-8}$ |
| chr11(q23.1) | 111609174 | rs647080 | G/A | 0.30 | 0.32 | 0.03 (0.005) | $2.8 \times 10^{-8}$ |
| chr13(q14.3) | 51159830 | rs947381 | T/C | 0.24 | 0.22 | 0.03 (0.005) | $2.1 \times 10^{-9}$ |
| chr13(q31.1) | 80572249 | rs9531009 | G/A | 0.27 | 0.27 | 0.03 (0.005) | $2.1 \times 10^{-8}$ |
| chr15(q15.1) | 40971835 | rs28866515 | T/A | 0.39 | 0.44 | 0.03 (0.004) | $9.1 \times 10^{-10}$ |
| chr17(q21.31) | 43170422 | rs2239920 | A/G | 0.39 | 0.45 | 0.02 (0.004) | $1.8 \times 10^{-8}$ |
| chr18(q21.32) | 57185289 | rs78560767 | C/T | 0.12 | 0.10 | 0.04 (0.007) | $3.6 \times 10^{-8}$ |
| chr20(p12.3) | 5817549 | rs237101 | A/G | 0.37 | 0.31 | 0.03 (0.005) | $4.3 \times 10^{-8}$ |

Further annotation for the novel loci revealed that four hypothyroidism loci (lead variants rs7520586, rs13238252, rs34053209, rs34988504) harbor genes (*CSF1*; *SNX10*, *HOXA1*, *HOXA3,* and *HOXA5*; *ELF1*; *MED1* and *THRA*) associated with thyroid phenotypes in mice (Figure 3; Supplementary table S4).

**Figure 3.**
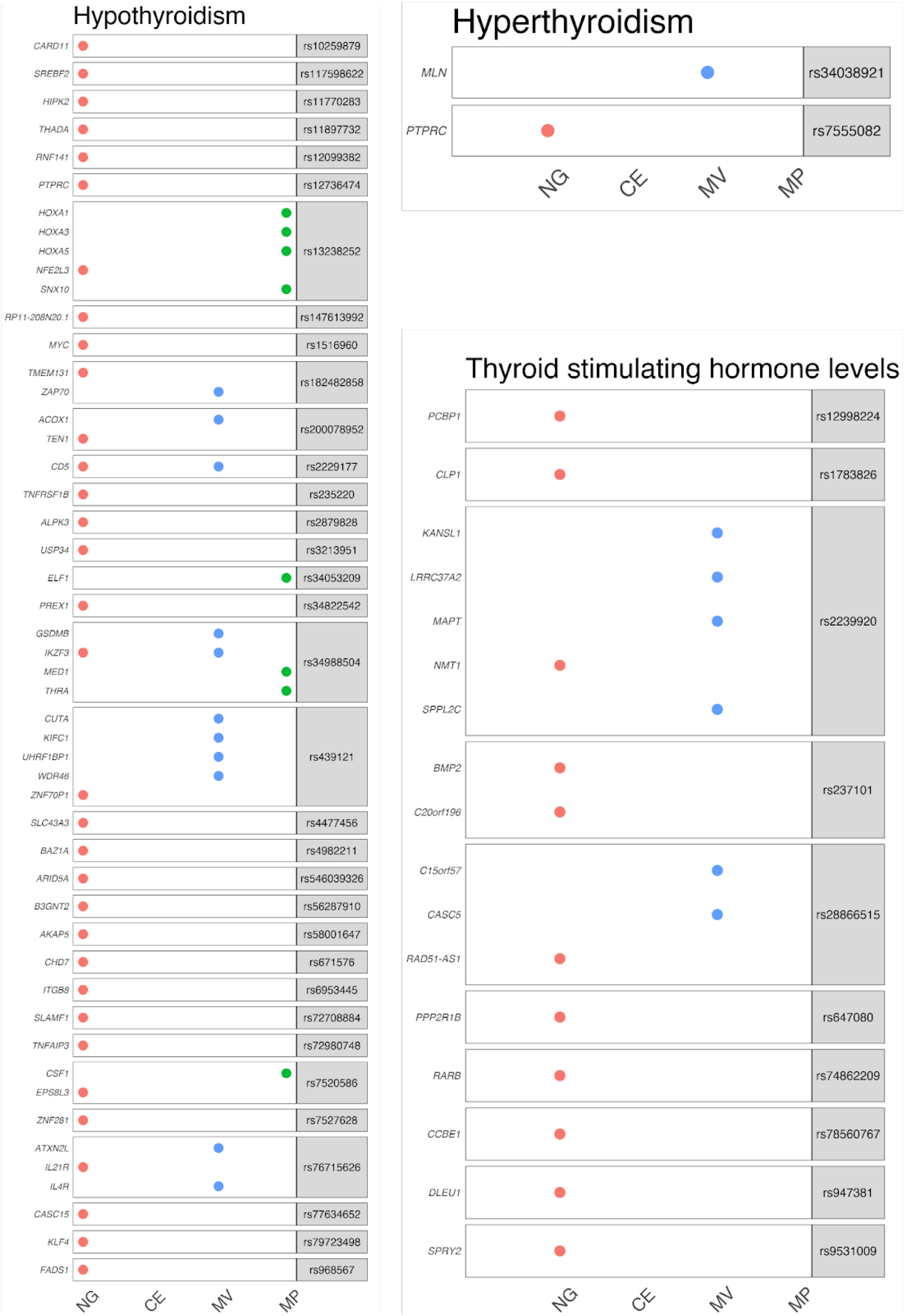
Gene prioritisation for novel variants of hyperthyroidism, hypothyroidism and TSH measurements levels. NG = nearest gene to lead signal; CE=colocalization evidence; MV= missense variants; MP= mouse phenotypes.

Overall, based on our parameters for gene prioritisation, 309 genes across both novel and previously reported loci presented at least one level of evidence of being linked with thyroid biology. Thus, they were considered as candidate genes for our associated loci (Figures 3 and 4; Supplementary table S4). Among those, three genes presented three levels of evidence: two genes from hypothyroidism (*TPO*, *TG*) and one gene from TSH levels analysis (*SOX9*). *SOX9* is involved in thyroid follicular cell differentiation, while *TPO* and *TG* are well-known genes responsible for synthesising and controlling thyroid hormones, and consequently play a role in autoimmune thyroid diseases (Citterio et al., 2019; Godlewska et al., 2019; López-Márquez et al., 2022; Mete et al., 2019).

**Figure 4.**
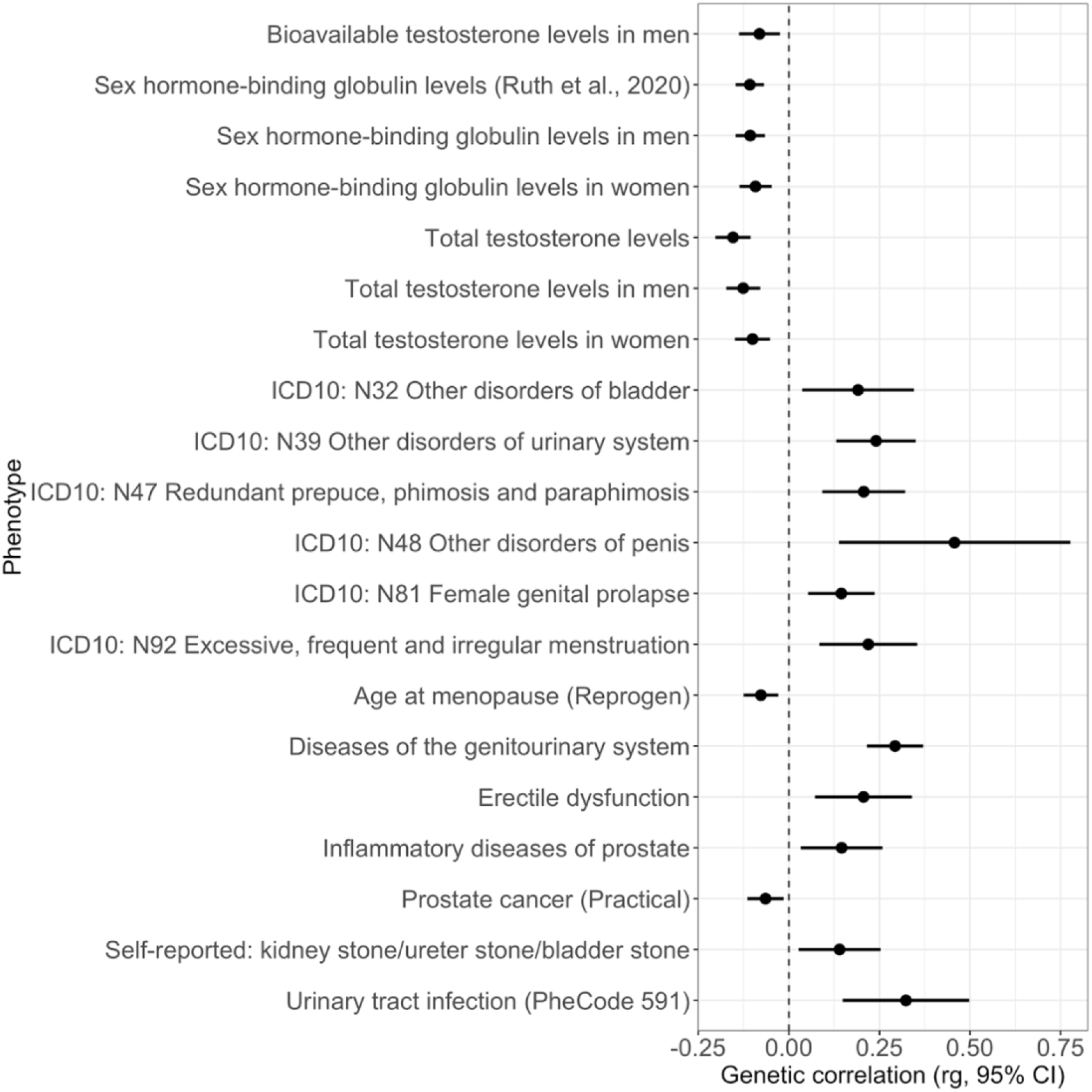
Significant (Benjamini-Hochberg FDR<0.05) genetic correlations between hypothyroidism and reproductive health phenotypes.

### Overlap between thyroid biology and reproductive health

We next explored the overlap between thyroid biology and reproductive health. For this, we used a multifaceted approach, analysing both phenotypic and genetic correlations and exploring the impact of candidate genes for thyroid phenotypes identified in the previous step.

To characterise the genetic variants, we conducted a search in the GWAS Catalog and literature. 32 out of 237 lead variants presented a link with both thyroid and reproductive health in previous literature. Considering the novel loci found in our study, seven of them showed association with reproductive health traits (rs2476491, rs11897732, rs56287910, rs13238252, rs2229177, rs968567, and rs647080). Most of the associations were related to female reproductive health and sex hormones, only the locus rs2229177 (prioritised gene *CD5*) showed specific association with male traits (prostate cancer) (Supplementary table S5).

According to the genetic correlation analysis, using the meta-analysis results and publicly available datasets, only hypothyroidism presented significant correlations with reproductive phenotypes after FDR correction for multiple testing. We identified a total of 23 correlations, nine related to sex hormone traits and 14 to reproductive traits. All significant genetic correlations are presented in Figure 4 and Supplementary table S6.

In addition to genetic correlations, we explored phenotypic associations in a Phe-PheWAS setting. Based on EstBB sex-stratified data, Phe-PheWAS results revealed 91 associations (p-value<0.05) with reproductive diagnoses. After applying the Bonferroni correction, 36 of these associations were statistically significant, of which 32 associations were specific to women and two were specific to men (p-value threshold ≤ 2.5 × 10^−5^) (Figure 5; Table 2; Supplementary figures SF1-SF2 and table S7). Furthermore, 44 of the at least nominally significant phenotypic associations overlapped with findings from the mouse phenotype lookup (Supplementary table S8).

**Figure 5.**
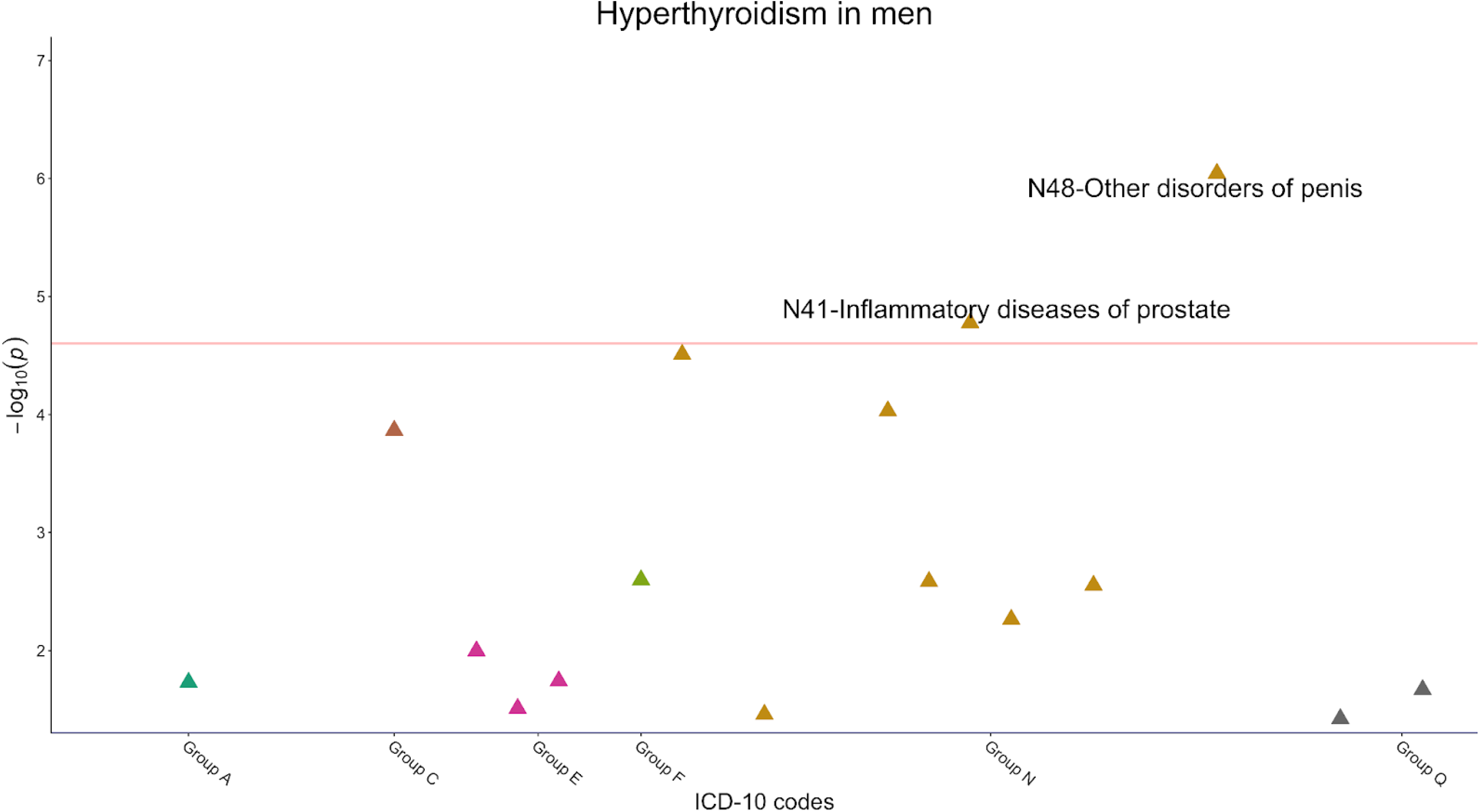
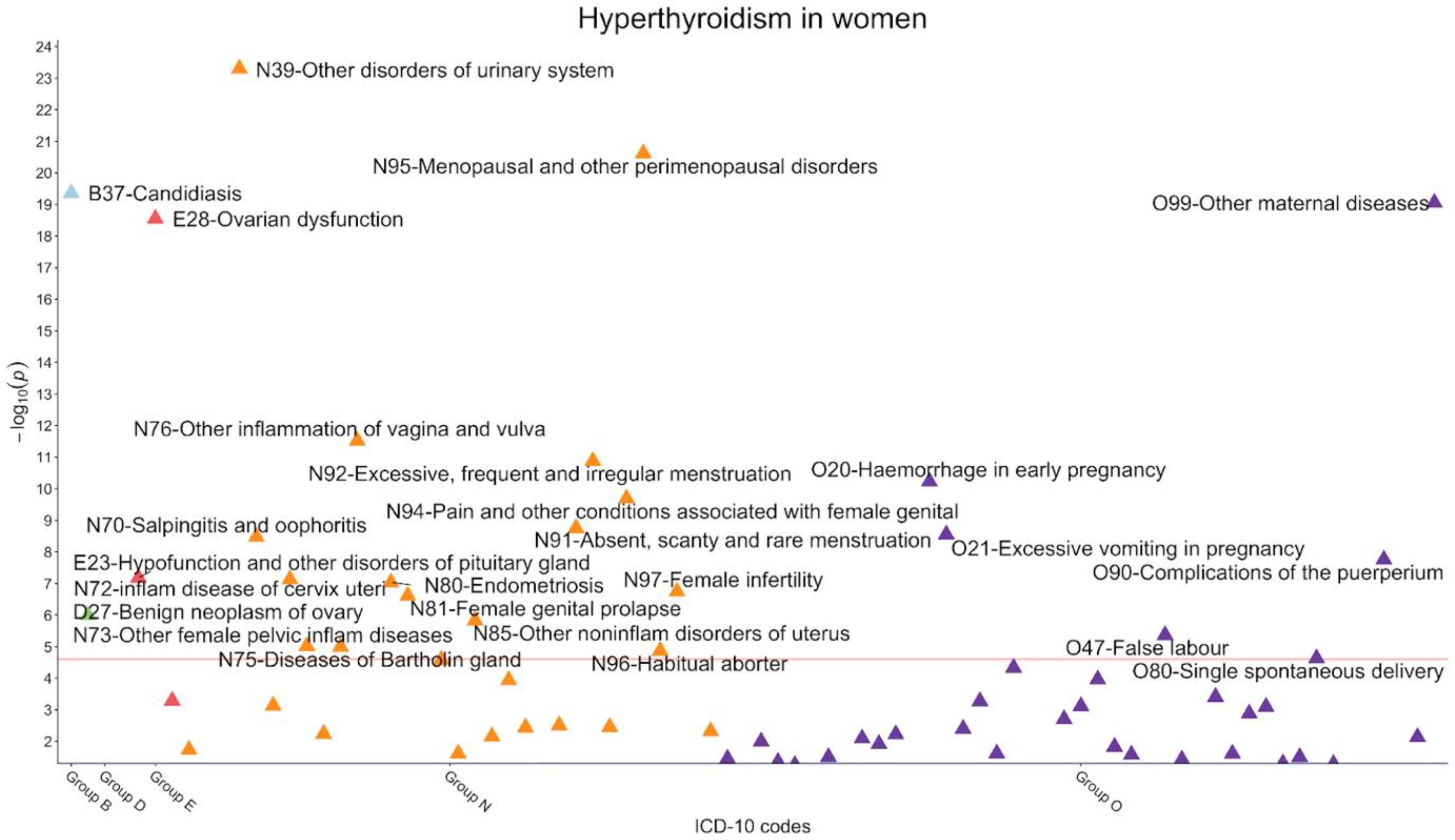
Phe-PheWAS results for EstBB individual level data for hyperthyroidism stratified for men and women. Only reproductive health related ICD-10 codes with p-value<0.05 are displayed in the figure, and the ones that passed Bonferroni correction are annotated. The red line stands for p-value threshold used in Bonferroni correction (p-value=2.50 x 10^−05^). Phe-PheWAS figures for hypothyroidism and TSH levels are in the supplementary files (Figures SF1 and SF2, respectively). Inflam= inflammatory.

**Table 2.**
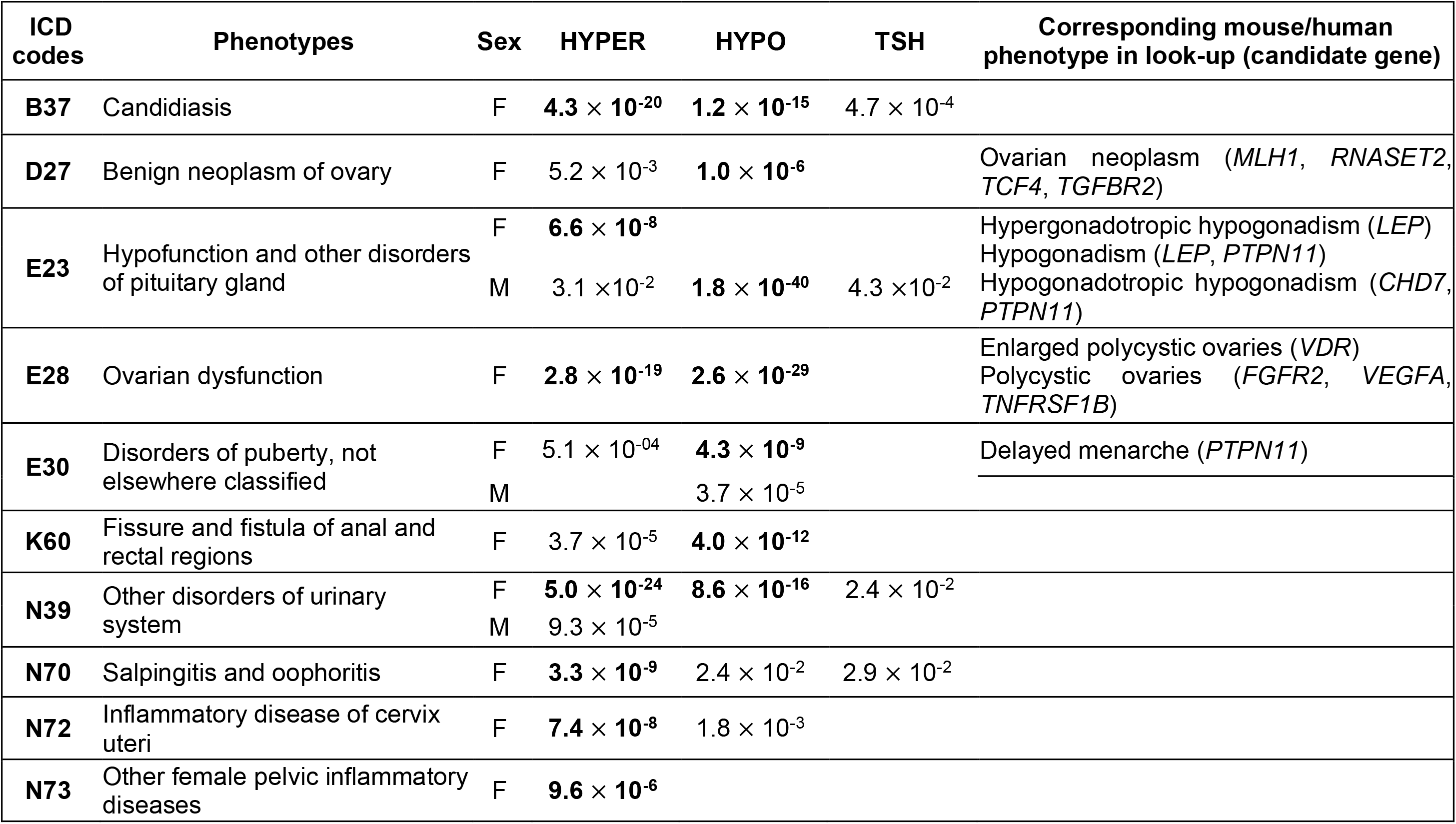
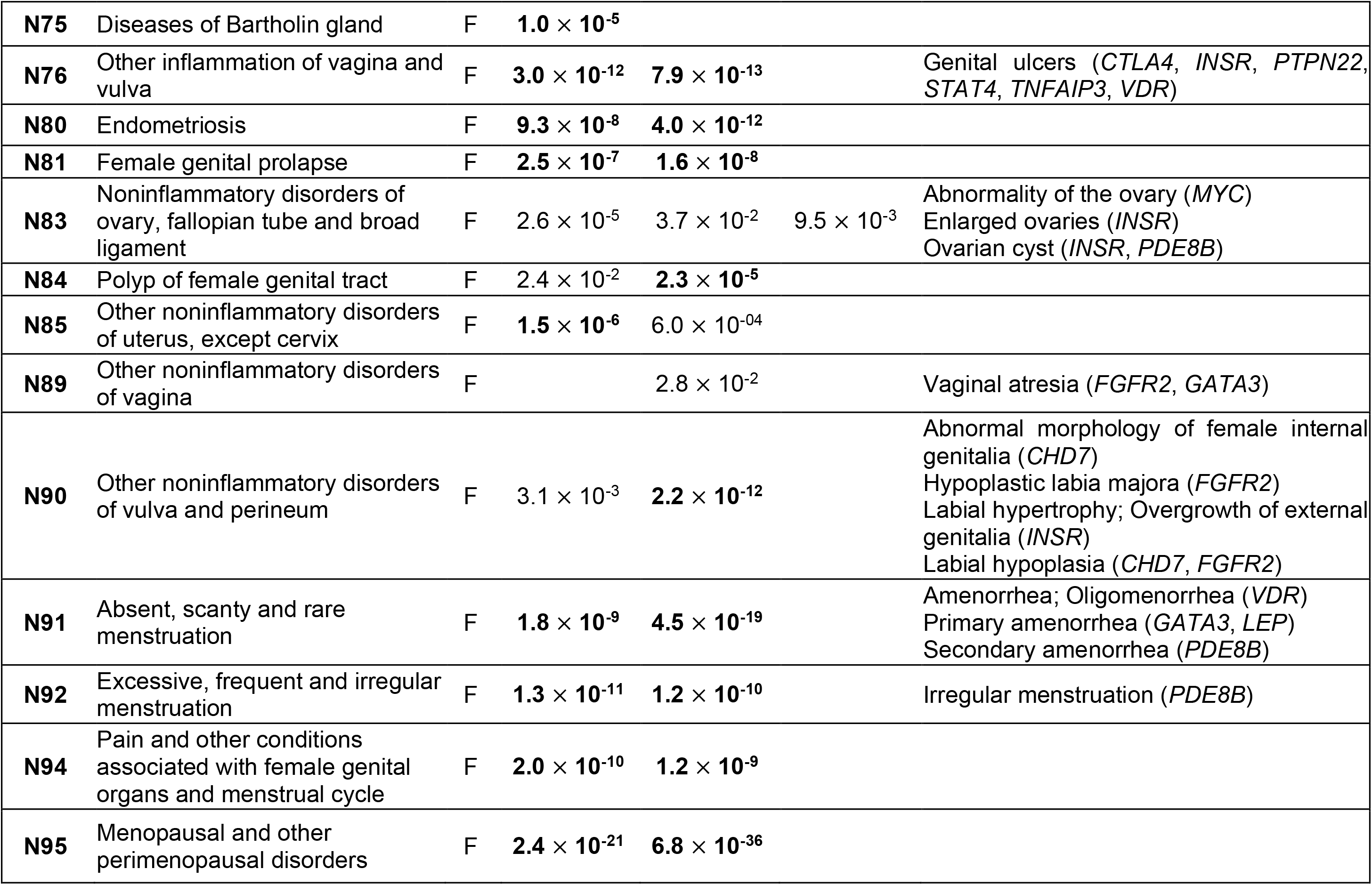
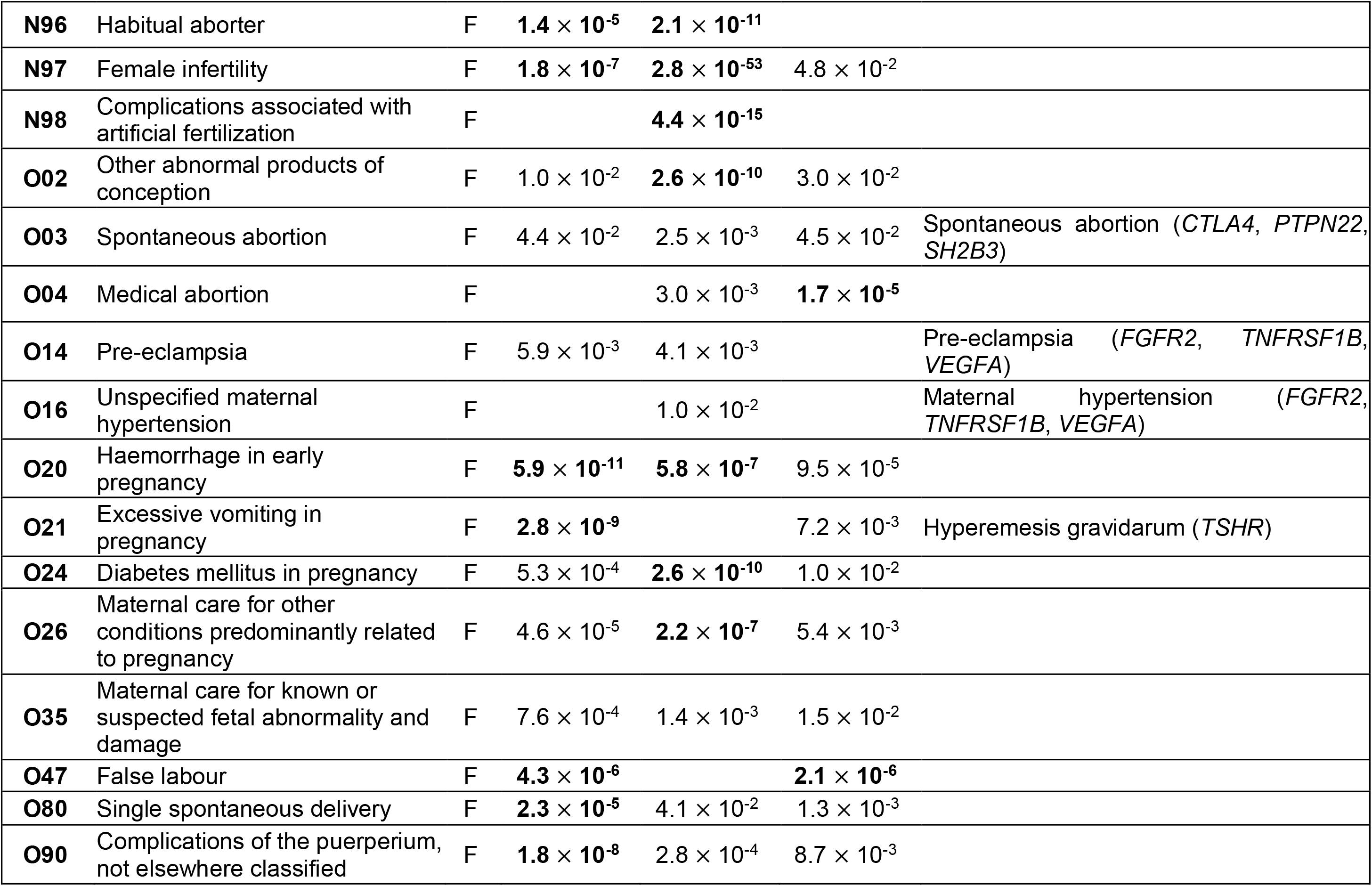
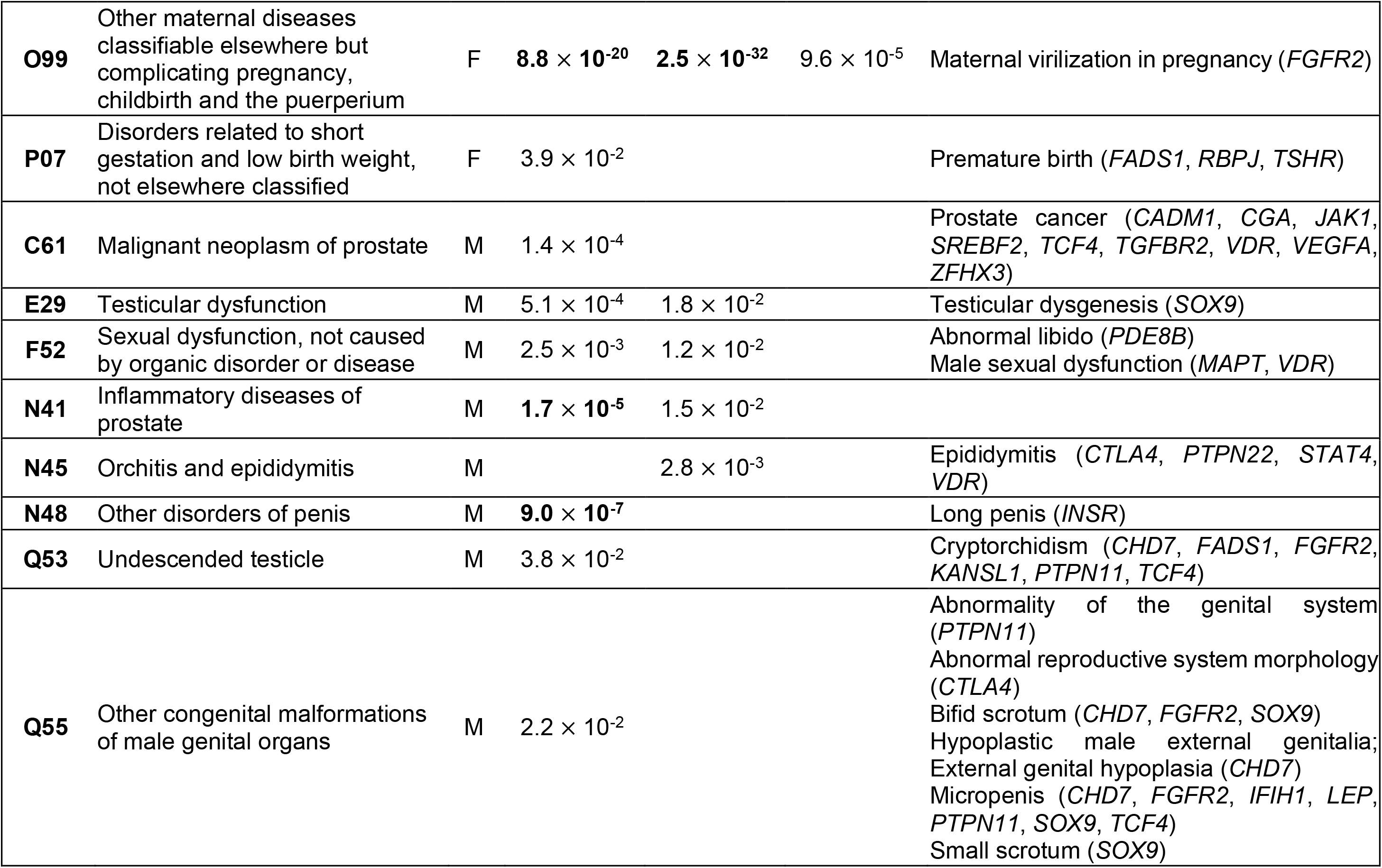
Phe-PheWas significant p-values for reproductive traits in female (F) and male (M), and corresponding human/mouse phenotype associated in look-up with their respective candidate genes. P-values statistically significant after Bonferroni correction are highlighted in **bold** (p-value threshold= 0.05/2001=2.5 x 10^−05^). ICD-10 codes followed version 2019.

Comparing the results from Phe-PheWAS and genetic correlation analysis, we found seven overlaps with specific diagnoses. For females we detected overlaps for: N32-Other disorders of bladder (Hypothyroidism-Phe-PheWAS p-value = 2.4 × 10^−3^, Genetic correlation p-value = 1.5 × 10^−2^), N39-Other disorders of urinary system (Hypothyroidism-Phe-PheWAS p-value = 8.6 × 10^−16^, Genetic correlation p-value = 1.8 × 10^−5^), N81-Female genital prolapse (Hypothyroidism-Phe-PheWAS p-value = 1.6 × 10^−8^, Genetic correlation p-value = 2 × 10^−3^), and N92-Excessive, frequent and irregular menstruation (Hypothyroidism-Phe-PheWAS p-value = 1.2 × 10^−10^, Genetic correlation p-value = 1.5 × 10^−3^). While for males we had overlap for: N41-Inflammatory diseases of prostate (Hypothyroidism-Phe-PheWAS p-value = 1.5 × 10^−2^, Genetic correlation p-value = 1.1 × 10^−2^), and for the phenotypes N48-Other disorders of penis and N39-Other disorders of urinary system showed association on genetic level with hypothyroidism (Genetic correlation p-values = 5 × 10^−3^ and 1.8 × 10^−5^, respectively) and on phenotypic level with hyperthyroidism (Hyperthyroidism-Phe-PheWAS p-values= 9.04 × 10^−7^ and 9.32 × 10^−5^, respectively).

Based on the lookup of mouse phenotypes, 99 prioritised genes are associated with any reproductive system phenotype, and 34 of them had associations with both reproductive system and thyroid phenotypes (Figure 6). Detailed information about the mouse phenotype associations is in Supplementary table S9.

**Figure 6.**
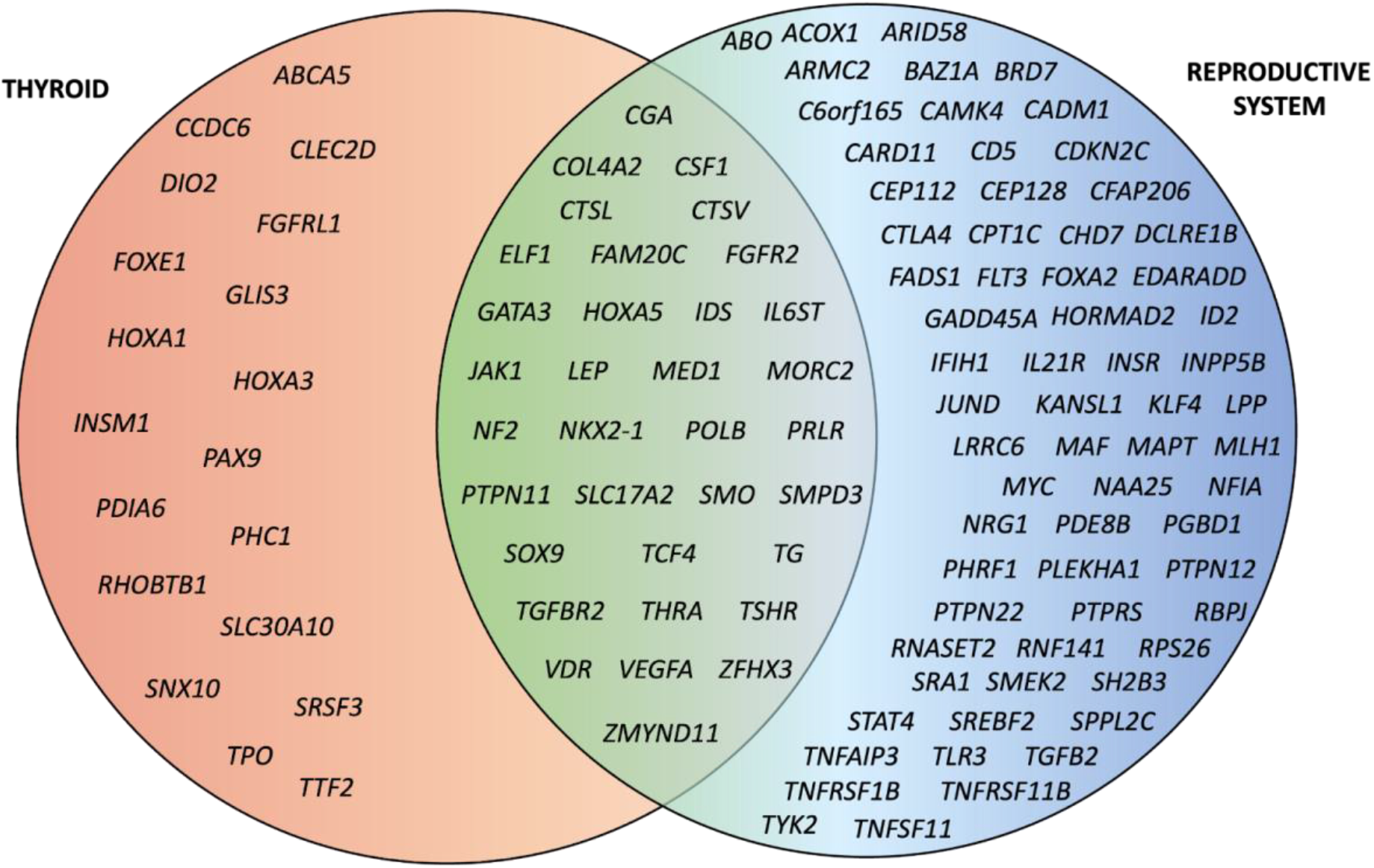
Overview of genes prioritised in thyroid analyses and their associations with thyroid and/or reproductive system according to mouse phenotype lookup.

To compare the findings between the mouse phenotype lookup and Phe-PheWAS analysis, we chose for reproductive traits equivalent ICD-10 codes based on the mouse phenotypes. In total, we had an overlap of 23 phenotypes, including 13 associations that did not pass the multiple testing correction in Phe-PheWAS analysis. Seven associations were present in men, 16 in women. One association (E23-Hypofunction and other disorders of pituitary gland) was present in both sexes and is relevant to the physiological axes for thyroid and sex hormone production. In sum, 41 genes associated with the overlapping mouse phenotypes, of which 16 (*CGA*, *FGFR2, GATA3*, *JAK1*, *LEP*, NF2, *POLB*, *PTPN11*, *SMO, SOX9, TCF4*, *TGFBR2*, *TSHR, VDR, VEGFA* and *ZFHX3*) also presented human/mouse phenotypic association with thyroid phenotypes (Table 2; Supplementary table S8).

As an interesting example, ‘Excessive, frequent and irregular menstruation’ (N92) was associated with thyroid traits in the three analyses: genetic correlation, Phe-Phewas and mouse/human phenotype look-up for candidate genes. In the last analysis, the phenotype was linked with the gene *PDE8B*, a likely candidate gene for hypothyroidism and TSH levels in the gene prioritisation analysis at chr5(q13.3), lead signals rs7727976 and rs6885099, respectively.

## DISCUSSION

This study describes the genetic and phenotypic association between three thyroid traits and reproductive health traits, using GWAS results of hyper- and hypothyroidism and TSH levels to map the shared genetic components. Using a multifaceted mapping approach combining different levels of evidence, we prioritise 309 genes in the thyroid analyses. Further analysis based on evidence from human conditions and mouse phenotypes revealed potential roles for 99 of these genes in reproductive health, providing evidence for pleiotropic effects. Our genetic correlation analyses also highlighted shared genetics between hypothyroidism, sex hormone levels and genitourinary tract disorders. Additionally, our phenotype-phenome wide association studies (Phe-PheWAS) analysis utilising individual level data from the Estonian Biobank, revealed 36 phenotypic associations between thyroid conditions and reproductive traits.

Three hypothyroidism lead variants (rs546039326, rs182482858, rs76715626) showed higher effect allele frequency in EstBB and FinnGen cohorts, when compared to other Europeans from UKBB (Chen et al., 2022). That reinforces the relevance of these population specific studies for expanding our understanding of disease biology (Tyrmi et al., 2022).

In general, our study finds that 32% of proposed thyroid-associated genes likely also affect genitourinary phenotypes, since out of the 309 candidate genes, 99 exhibited a reproductive system phenotype in either mouse or humans. Various genital tract issues (such as female genital prolapse, erectile dysfunction, prostate cancer, urinary tract infection and other disorders of bladder and urinary system) were the most prevalent. In line with previous reports (Krassas and Markou, 2019), we also found associations with menstrual irregularities, which was evident in every analysis performed.

The genetic correlation assessment provided evidence of significant associations between thyroid disorders and reproductive hormones and diseases. Through the positive association between TSH and HNP-4α in the liver, thyroid hormones can indirectly increase the SHBG production (Selva and Hammond, 2009). The main function of SHBG is the transport of androgens (e.g. androstenedione, DHEA and T) and estrogens (e.g. estradiol, estrone and estriol) in the blood and regulating the bioavailability of those hormones in target tissues. Considering this, thyroid dysfunction can have a relevant impact on the concentrations of reproductive hormones in the body, meaning that the concentration of the thyroid hormones has a positive correlation with reproductive hormones (Song et al., 2015). In line with previous findings (Kjaergaard et al., 2021; Krassas and Markou, 2019), hypothyroidism had a negative genetic correlation with reproductive hormones (SHBG, total and bioavailable T levels), since hypothyroidism is a consequence of a low thyroid hormone concentration in the blood.

Our study shows a positive phenotypic and genetic association between thyroid diseases, female genital prolapse, and disorders of the bladder/urinary system. Individuals with thyroid problems often present lower urinary tract symptoms (Zargham et al., 2022). Although the topic has not been studied thoroughly and the exact mechanisms remain unclear, it is known that thyroid hormones also affect the gastrointestinal tract and kidney function (Frau et al., 2017; Iglesias et al., 2017). Whether the link with prolapse stems from symptoms shared between prolapse and thyroid-related gastrointestinal tract and renal issues, which can result in initial misdiagnosis, or there is some other mechanism involved, remains to be determined.

Thyroid traits showed association with ’Excessive, frequent and irregular menstruation’ (N92) and link with the gene *PDE8B* in the mouse/human phenotype look-up. *PDE8B* is associated with thyroid-hormone levels and affects the course of TSH release by the pituitary in humans (Arnaud-Lopez et al., 2008). The association between menstrual cycle disturbances and endocrine diseases has broadly been discussed in the literature (Naz et al., 2020), and our study confirms the association also on a genetic level.

The positive genetic correlation between hypothyroidism and erectile dysfunction supports the idea that thyroid hormones are involved with sexual functioning. Studies have reported this association and it is recommended screening for endocrine diseases in patients with sexual dysfunction (Chen et al., 2018; Nikoobakht et al., 2012). Besides the abovementioned interaction between HPT and HPG axes, we provide further evidence that the *INSR* gene may also be associated with both thyroid function and erectile dysfunction. We identified *INSR* as a potential causal gene at a locus associated with TSH levels (lead signal rs12610987). *INSR* has been associated with erectile dysfunction previously (Kazemi et al., 2021), with other genital tract issues in humans (like labial and clitoral hypertrophy, long penis, overgrowth of external genitalia, enlarged ovaries and ovarian cyst) and also in both male and female mice (oligozoospermia, decreased corpora lutea number, decreased tertiary ovarian follicle number, abnormal seminiferous tubule morphology, decreased Leydig cell number, abnormal spermatogenesis, reduced male fertility).

Although our study brings proof of shared genetics between thyroid phenotypes and reproductive health, there are some limitations that need to be considered when interpreting our results. First, thyroid disorders may have pathophysiologically different subphenotypes, which were grouped together in our analyses to gain power and due to pre-existing phenotype definitions in publicly available GWAS browsers. Second, all the cohorts included in our analyses are of European ancestry, meaning caution is needed when extrapolating the results to non-European populations. Multi-ancestry analyses are needed to explore the genetic intersection of thyroid and reproductive biology in other ancestries. Third, our selected study design only shows correlations and associations but not causality.

In conclusion, our study provides a roadmap to understand the shared genetics and genetic pleiotropy between thyroid function and reproductive health, both in men and women. The identified phenotypic and genetic correlations provide valuable insights into the potential comorbidities of thyroid problems, however studies evaluating the causality and direction of the observed associations are needed.

## Data Availability

All data produced in the present study are available upon reasonable request to the authors.

## Acknowledgements

This study was funded by the European Union through the European Regional Development Fund Project No. 2014-2020.4.01.15-0012 GENTRANSMED, and by the Estonian Research Council grants 1911, PRG1844 and PRG1291.

The activities of the EstBB are regulated by the Human Genes Research Act, which was adopted in 2000 specifically for the operations of the EstBB. Individual level data analysis in the EstBB was carried out under ethical approval 1.1-12/624 from the Estonian Committee on Bioethics and Human Research (Estonian Ministry of Social Affairs), using data according to release application S53 from the Estonian Biobank.

Data analysis was carried out in part in the High-Performance Computing Center of University of Tartu. We want to acknowledge the participants and investigators of the FinnGen study. Natàlia Pujol-Gualdo was supported by MATER Marie Sklodowska-Curie which received funding from the European Union’s Horizon 2020 research and innovation program under grant agreement No. 813707.

The Genotype-Tissue Expression (GTEx) Project was supported by the Common Fund of the Office of the Director of the National Institutes of Health, and by NCI, NHGRI, NHLBI, NIDA, NIMH, and NINDS. The data used for the analyses described in this manuscript were obtained from eQTL Catalog on 05/25/2023.

