## Supplementary material for "Uncovering the Shared Genetic Components of Thyroid Disorders and Reproductive Health": SF1-SF2

**Supplementary Figure SF1.** Phe-PheWAS results for Estonian biobank individual level data for hypothyroidism stratified for men and women. Only reproductive health related International Classification of Disease 10 (ICD-10) codes are displayed in the figure, and the ones that passed Bonferroni correction are annotated. The purple line stands for p-value = 0.05, and the red line stands for p-value threshold used in Bonferroni correction (p-value = $2.5 \times 10^{-5}$).

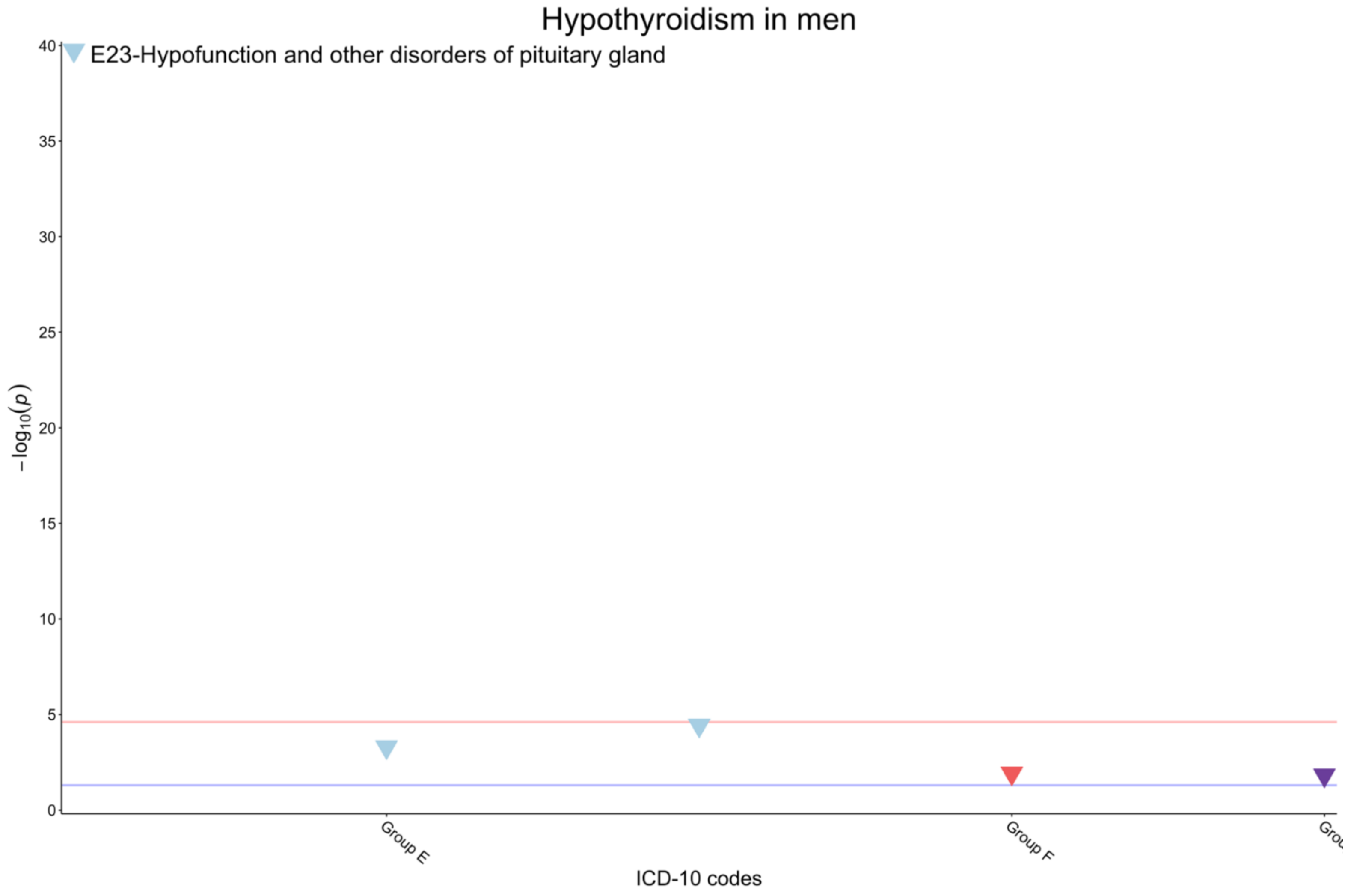

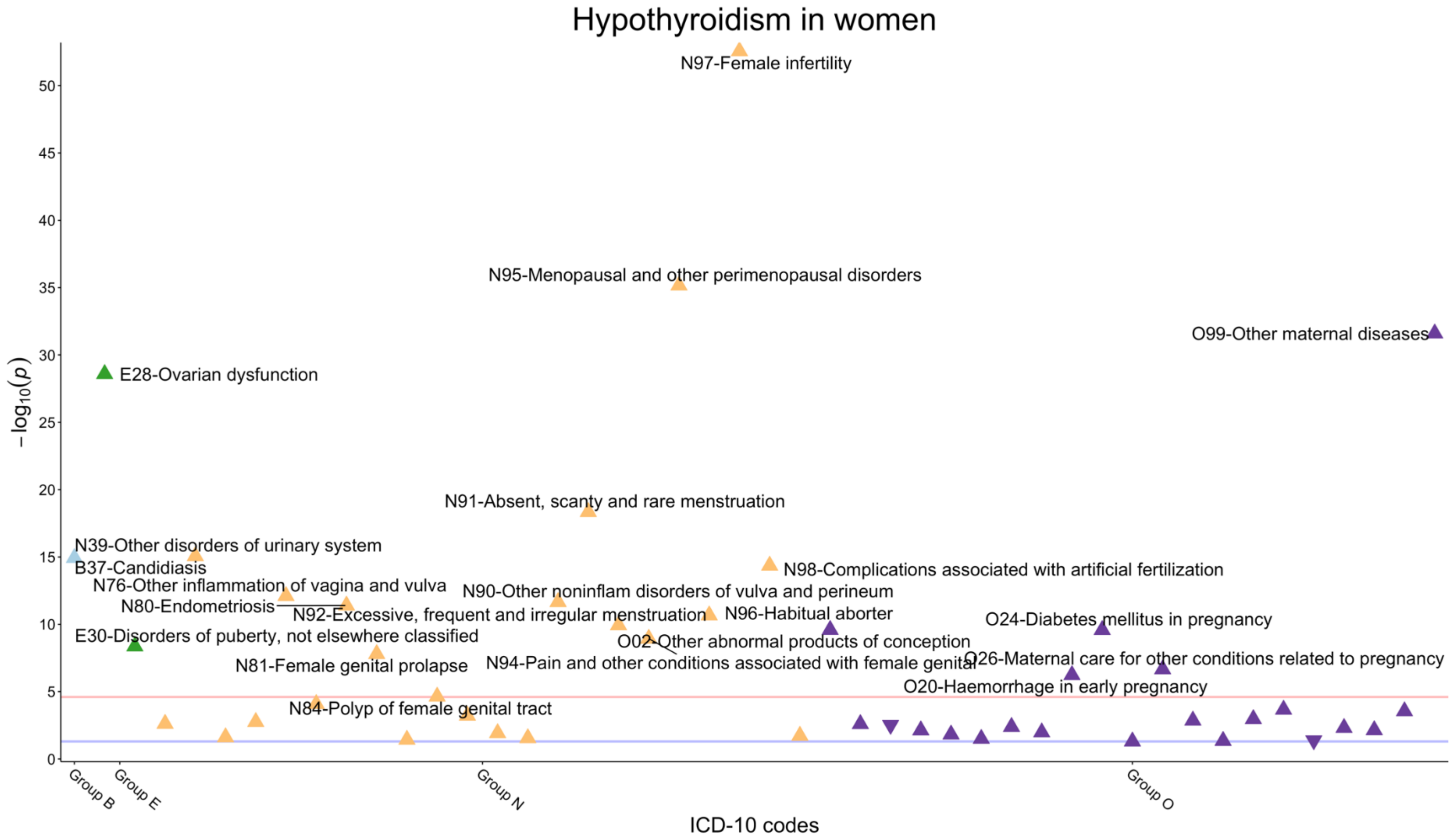

Hypothyroidism in women
N97-Female infertility
N95-Menopausal and other perimenopausal disorders
O99-Other maternal diseases
E28-Ovarian dysfunction
N91-Absent, scanty and rare menstruation
N39-Other disorders of urinary system
B37-Candidiasis
N98-Complications associated with artificial fertilization
N76-Other inflammation of vagina and vulva
N90-Other noninflam disorders of vulva and perineum
N80-Endometriosis
N92-Excessive, frequent and irregular menstruation
N96-Habitual aborter
O24-Diabetes mellitus in pregnancy
E30-Disorders of puberty, not elsewhere classified
O02-Other abnormal products of conception
O26-Maternal care for other conditions related to pregnancy
N81-Female genital prolapse
N94-Pain and other conditions associated with female genital
O20-Haemorrhage in early pregnancy
N84-Polyp of female genital tract
$-\log_{10}(p)$
0
5
10
15
20
25
30
35
40
45
50
Group B
Group E
Group N
Group O
ICD-10 codes

**Supplementary Figure SF2.** Phe-PheWAS results for EstBB individual level data for thyroid-stimulating hormone (TSH) levels stratified for men and women. Only reproductive health related ICD-10 codes are displayed in the figure, and the ones that passed Bonferroni correction are annotated. The purple line stands for p-value = 0.05, and the red line stands for p-value threshold used in Bonferroni correction (p-value = 2.5 x $10^{-5}$).

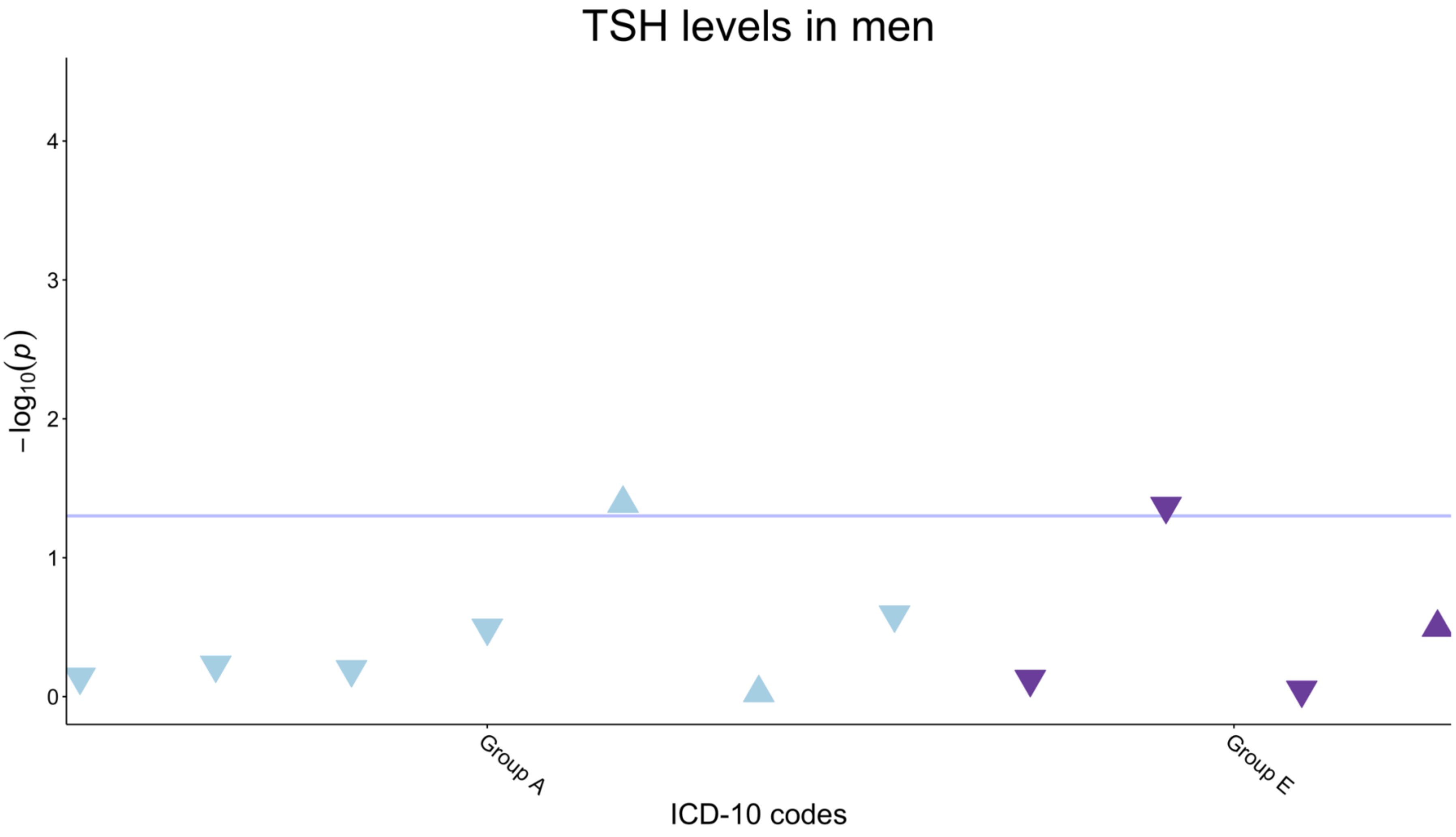

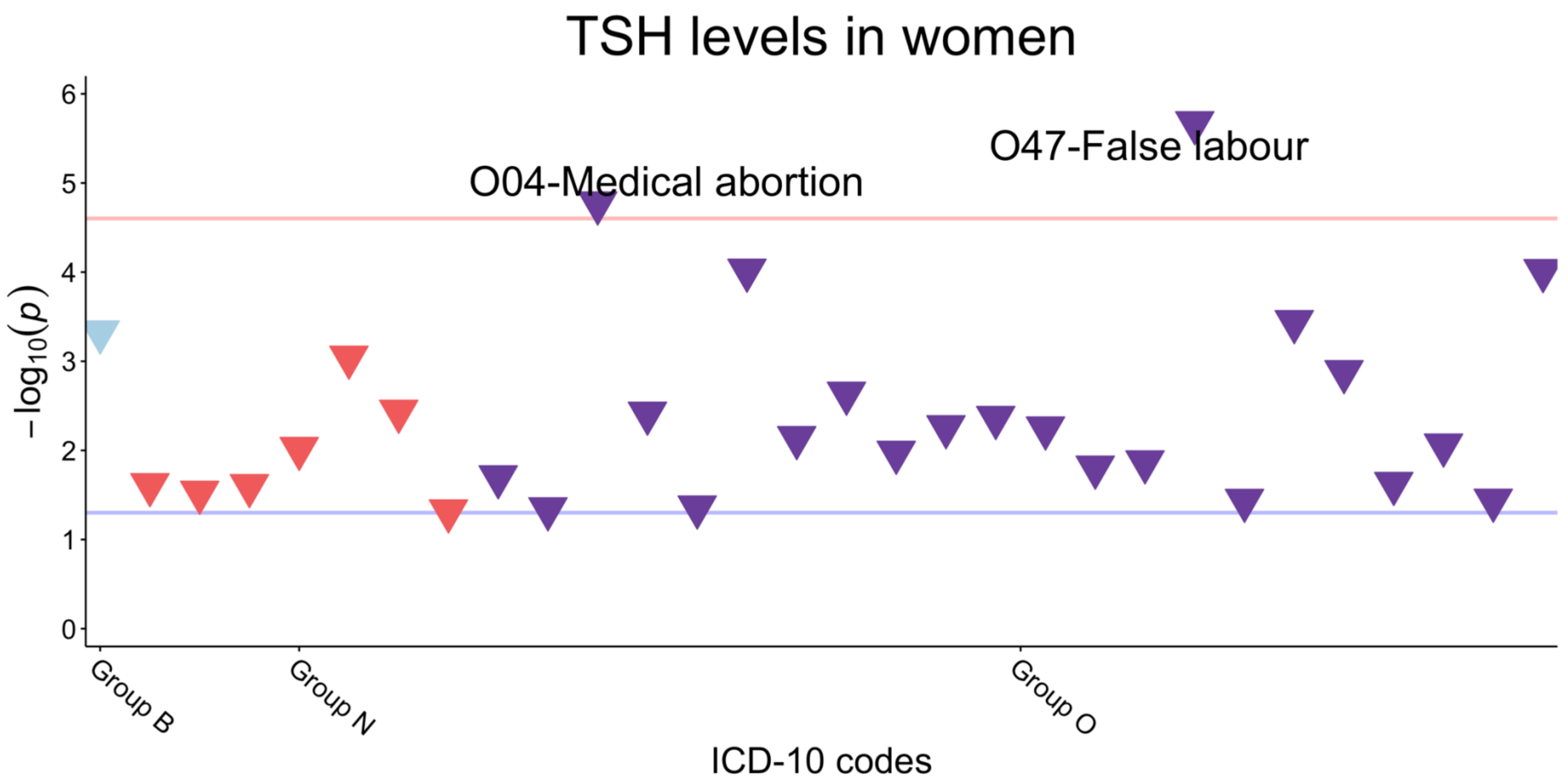

TSH levels in women
O47-False labour
O04-Medical abortion
6
5
4
3
2
1
0
$-\log_{10}(p)$
Group B
Group N
Group O
ICD-10 codes
